# HEALTH ECONOMIC BURDEN ON HEPATITIS B IMMUNOGLOBULIN VACCINATION

**DOI:** 10.1101/2022.06.11.22276274

**Authors:** Arbeen A. Laurito, Lourdes L. Bett

**Affiliations:** JR Borja General Hospital, Cagayan De Oro City, Philippines; Xavier University- Ateneo De Cagayan, Senior High School-Faculty

## Abstract

**Background:** Chronic Hepatitis B infection comprises the mortality among viral hepatitis despite primary hepatitis B vaccination was implemented in different states health programs. Different modalities of combining active and passive hepatitis B vaccination were conducted. Pregnant women hepatitis screening was not yet streamlined in the clinical management due to social and economic challenges. Fetal to maternal vertical transmission of hepatitis B virus is still a burden to our health system. It is the objective of the study to present the cost of incorporating hepatitis B immunoglobulin vaccine in the current vaccination program, as strengthen the preventive measures of hepatitis infection locally and nationally.

**Methods:** Survey questionnaires were utilized to gather demographic data among randomly selected pregnant women during their prenatal visits at the hospital. Hospital and city-based census were used for projecting cost and revenues of having hepatitis B immunoglobulin vaccination.

**Results:** A total of 74 respondents were identified. A financial 5-year was forecast would show a revenue of Php 1,494,500.00 ($28,869.5) on the 5^th^ year and would spare the mothers from OOP expenses with a total amount of Php 945,500 ($18,264.40). Moreover, the monthly revenue of Hep B Ig vaccination (based on 2016 census) on a city-wide was forecasted. The difference of the total amount of PHIC reimbursement from the amount of vaccine purchasing would give the facility a projected revenue of Php 3,381,123.00 ($65,313.62) or a monthly average of Php (281,760.25). A total of 1,380 newborns would be at risk to hepatitis B-reactive mothers and would be protected by securing hepatitis B immunoglobulin and be made available in the hospital pharmacy. Furthermore, pregnant women are protected against financial risk of unnecessary out-of-pocket expenses.

**Conclusion:** This study found that the pregnant women was aware of the economic burden of hepatitis B immunoglobulin vaccine and it would benefit the healthcare facility by strategically addressing the external factors for a sustained vaccination program.

## Introduction

Hep B Ig is not covered in the government’s Expanded Program of Immunization (EPI) program of vaccination. But once the newborn is exposed to a Hepatitis B-confirmed mother, Hep B Ig is time-bound and highly recommended to be given in less than 12 hours for better efficacy and protection against adult hepatitis B infection. Infection in infancy and early childhood leads to chronic hepatitis in about 95% of cases^**1**^. This is the basis for strengthening and prioritizing infant and childhood vaccination including Hepatitis B vaccine among all newborns in the Philippines.

World Health Organization (WHO) estimates that 296 million people were living with chronic hepatitis B infection in 2019, with 1.5 million new infections each year. The burden of hepatitis B infection is highest in the WHO Western Pacific Region and the WHO African Region, where 116 million and 81 million people, respectively, are chronically infected. In 2019, hepatitis B resulted in an estimated 820 000 deaths, mostly from cirrhosis and hepatocellular carcinoma (primary liver cancer)^**1**.^ In highly endemic areas, hepatitis B is most commonly spread from mother to child at birth (perinatal transmission) or through horizontal transmission (exposure to infected blood), especially from an infected child to an uninfected child during the first 5 years of life.

The health facility where the study was conducted had an average of 3,931 OB cases with newborns of 2,185 (based on 2017 unpublished hospital data). Moreover, the serology logbook of the hospital’s laboratory documented an average of 40 HBsAG reactive mothers, among the other indications for a high-risk newborn for hepatitis B infection. The hospital is a government-owned health facility and the apex facility which caters to the increasing city population with a predicted growth rate of 2.23%^**2**^.

Currently, the healthcare facility does not purchase the vaccine due to its cost and unstable target population. In this situation, the mother or relatives have to spend their own money for the Hep B Ig vaccine in the nearest private hospital, pharmacy or clinics. They can also avail lower cost of the vaccines in pharmacies outside a tertiary hospital but the distance is a challenge. Fortunately, stocks are available most of the time.

Thus, it is the objective of the study to determine vaccine access in the community, the preference of the mothers for the Hep B Ig vaccine, and its projected cost when the health facility planned to purchase the vaccine. This study will also give us a good and logical data on the burden of Hep B Ig cost from the perspectives of an insured mother from the national health insurance (Philippine Health Insurance Corporation or PHIC for brevity), and the health facility procurement program. Furthermore, it will help in reducing the out-of-pocket expenses among the target population and increases health equities.

### Logical Framework of the Study

The logic model was used in this study. The model clarifies who will be the target population to receive the input of the program, expected activity to achieve accomplishments, and the impact of this program prospectively. This particular study will be the baseline and the input to this program to contribute in reducing chronic hepatitis B infection and the protection against out-of-pocket expenses. Expected output, outcome and impact of the program are presented in Figure 1. But these are beyond the scope of the study.

**Figure 1.**
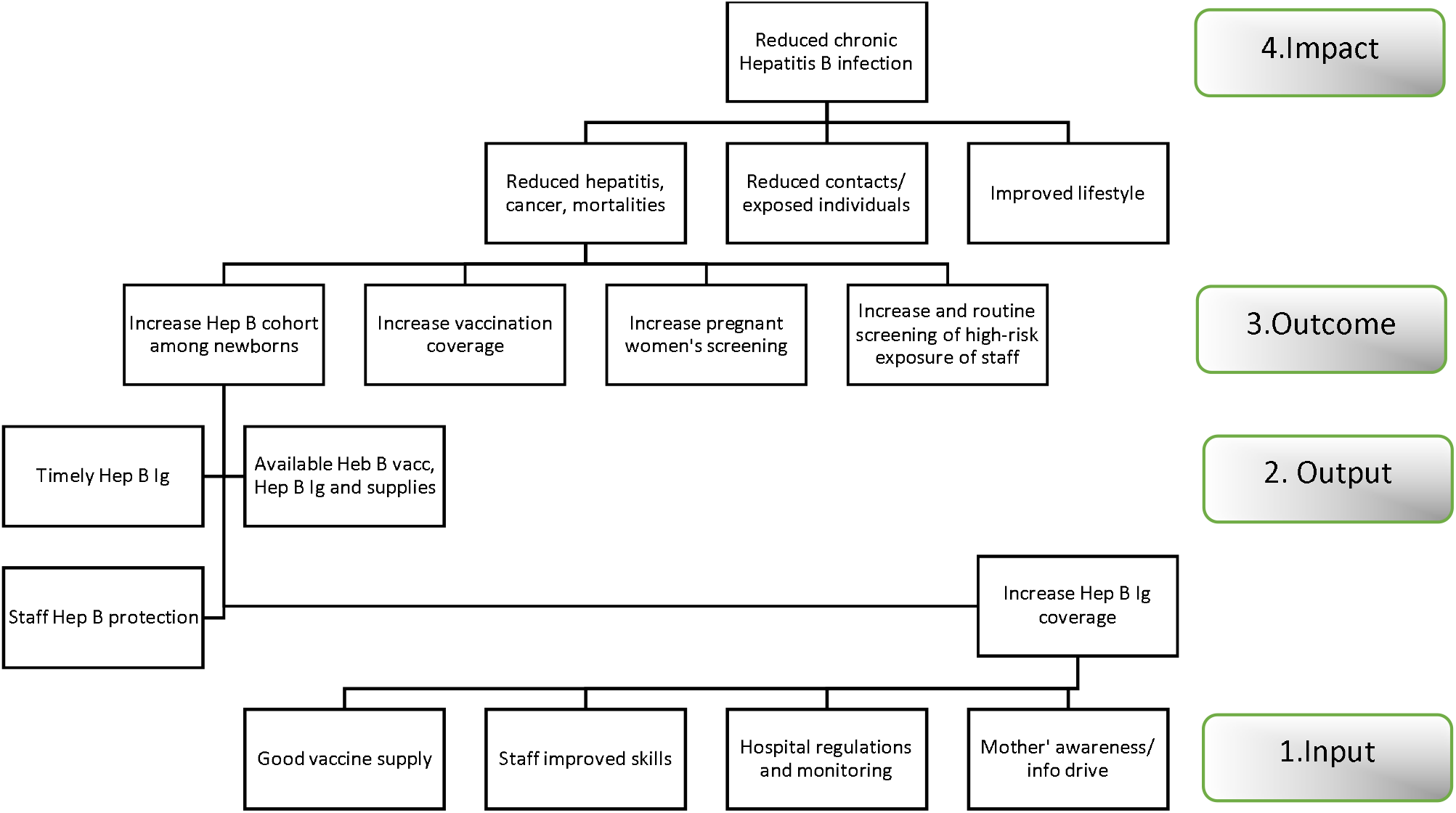
Schematic diagram of Hep B Ig vaccination based on Logic Model.

## METHODS

### Health economic analysis plan

This study planned to present the projected 5-year revenue based on admitted obstetrics cases in the hospital, projected 5-year revenue based on the city-wide population and use of questionnaire surveys among the respondents.

### Study population

Pregnant women who had their prenatal care (PNC) attended the family planning clinic at the Out-patient department of the hospital. Informed consent was discussed and secured from each respondent before the introduction of the questionnaire survey forms.

### Setting and location

Out-patient department of the level-1 public hospital J.R. Borja General Hospital (JRBGH) is located at Cagayan De Oro City, Misamis Oriental, Philippines. The city has 80 barangays clustered into 4 groups.

### Comparators

The current clinical management of a newborn born to a Hepatitis B reactive mother is to give a Hep B Immunoglobulin (Hep B Ig) within 12hours after birth^3,4^ for the optimal protection of the newborn. Since Hep B Ig is recommended but not readily available in the local setting, situational analysis was performed. Survey was done for the vaccine acceptance of the end-user and SWOT analysis for a comprehensive viewpoint for the needs of accessing Hep B Ig in each healthcare facility

### Perspective

Hep B Ig is critically important to be available anytime since there is a 12-hour golden period for its optimal effectiveness and should be accessible and affordable 24 hours and 7 days a week. Delayed giving of Hep B Ig to newborns via intramuscular route increases the probability of hepatitis infection in the adult age.

### Time horizon

The awareness of pregnant women is expectedly higher for her pregnancy and her baby’s health. Appropriate and good planning before delivery of an expectant of pregnant women is best during the prenatal period visits. Therefore, this study planned to determine the level of awareness and the capability of the pregnant women to proactively plan forward for her baby’s hep B Ig vaccination, if applicable.

### Selection of outcomes

Demographic profiles of the respondents were gathered through the questionnaire surveys, determined their knowledge and preference relative to hep B Ig vaccination, SWOT analysis for the needs of sustaining the program, forecast revenues of the health facility based on assumptions in a 5-year time frame.

### Measurement of outcomes

The survey answers were the bases for the values to be utilized in the 5-year forecast of Hep Ig vaccination costs, collection of admission census from the health facility, and the prevailing cost in dollars.

### Analytics and assumptions

Descriptive analyses were presented in terms of frequency, mean, mode and percentage on the demographic profile of the respondents and their knowledge on Hep B Ig vaccination. The following assumptions considered were as follows; Assumptions A -The forecast assumptions are based on the JRBGH Obstetric cases who were confirmed as Hep B-reactive mothers with their babies, and Assumptions B-The forecast assumptions were based on the city-wide obstetrics monthly cases^**2**^.

## RESULTS

### A. Demographic Profile of Pregnant Women

Pregnant women who had their prenatal care (PNC) visits at JRBGH and attended the family planning clinic were the respondents of this survey questionnaire (n=74). The majority of respondents belong to the mature group of pregnant women with age ranges 25-35 years of age (40.7%), followed by age 19-24 years old (30.5%). There were 9 (15.2%) women aged more than 36 years of age while 8 teenage pregnant women (13.6%). The youngest among the respondents was 16 years old (Table 1).

**Table 1.** Demographic profile of pregnant women surveyed (n=74).

| Profile | No. | % | Profile | No. | % |
| --- | --- | --- | --- | --- | --- |
| **Age** | **59** |  | **Religion** | **73** |  |
| <19yo | 8 | 13.6 | Catholic | 56 | 76.7 |
| 19-24 yo | 18 | 30.5 | Protestants | 1 | 1.4 |
| 25-35 yo | 24 | 40.7 | Mormons | 1 | 1.4 |
| >36 yo | 9 | 15.2 | Others | 15 | 20.5 |
| **Marital Status** | **72** |  | **Income** | **71** |  |
| Married | 24 | 33.3 | <5,000 | 42 | 59.1 |
| Live-in | 34 | 47.2 | 5,000-10,000 | 26 | 36.6 |
| Single | 14 | 19.4 | 10,000-20,000 | 2 | 2.8 |
|  |  |  | >20,000 | 1 | 1.4 |
| **Children** | **47** |  | **Educational level** | **72** |  |
| Living | 41 | 87.2 | Elementary level | 3 | 4.2 |
| Fullterm | 3 | 6.4 | Elementary Grad | 2 | 2.8 |
| Preterm>5 | 0 | 0 | High School level | 17 | 23.6 |
| Abort | 3 | 6.4 | High School Grad | 25 | 34.7 |
|  |  |  | Senior HS | 2 | 2.8 |
|  |  |  | College | 23 | 31.9 |

Most of these respondents were Roman Catholics, 56 (76.7%), living with their partners, 34 (47.2%), and with a monthly income bracket below Php 5,000 (59.1%). Though we have 8 teenage pregnant respondents, they attributed to the majority with living children, 41 (87.2%), which means most of them are multigravida (1 or more pregnancy). The mode of distribution of the respondents based on their educational level belongs to the High School level or graduate.

The addresses of the respondents’ residents were well distributed all throughout the city and the neighboring municipalities. Table 2 showed the distribution of the respondents representing the top ten most populated barangays. The other 34 (48.6%) respondents came from other barangays of the city with 1-3 respondents each. Two (2) of the respondents did not indicate their address.

**Table 2.** Distribution of the respondent based on the top 10 barangays of the city.

| Barangay | Population | Respondent |
| --- | --- | --- |
| Carmen | 70,492 | **10** |
| Lapasan | 43,611 | **3** |
| Kauswagan | 35,069 | **5** |
| Balulang | 34,793 | **3** |
| Bulua | 32,348 | **5** |
| Camaman-an | 30,927 | **1** |
| Bugo | 30,893 | **3** |
| Canito-an | 27,815 | **2** |
| Gusa | 26,815 | **5** |
| Iponan | 26,340 | **1** |
| Total | 359,103 | **38** |

Note: N=38 + 36 (other barangays) = 74

JRBGH is the referring health facility for these barangay health centers. Therefore, the majority of these respondents were referred from medical staff from these health centers their areas (42, 57.5%); some by their relatives, 10 (13.7%); and some visited based on their personal preference, 16 (21.9%) as seen in Table 3.

**Table 3.** Prenatal care status of pregnant women surveyed (n=74).

| Reasons for JRBGH referral | No. | % | PNC Done at | No. | % |
| --- | --- | --- | --- | --- | --- |
| Referral from relatives | **10** | **13.7** | BHC | **43** | **58.9** |
| Referral from the medical staff | **42** | **57.5** | JRBGH | **28** | **38.4** |
| Personal preference | **16** | **21.9** | Private hospital | **2** | **2.4** |
| Others | **5** | **6.8** | Private MD | **0** | **0** |
|  |  |  | Others | **0** | **0** |
|  |  |  | None | **0** | **0** |
| Total | **73** | **100** |  | **73** | **100** |

Furthermore, 43 (58.9%) of the respondents had their initial PNC at the barangay health centers, and some were referred due to indications of high-risk pregnancies. About 38.4% (28) had their first PNC at the hospital and 2 (2.4%) pregnant women had their initial PNC from a private hospital.

To assess the knowledge of these respondents, Table 4 showed that majority of them had previous history of vaccination (86.3%); mothers were willing to have vaccine injection (92.9%); mothers were willing to have their baby vaccinated (98.6%); Hep B Ig vaccination was explained to them (56.9%); and they were willing to have Hep B Ig vaccination for their babies if they were screened to be HBsAg reactive (90.4%). This will have a positive impact on the hep B Ig compliance due to high vaccine acceptance.

**Table 4.**
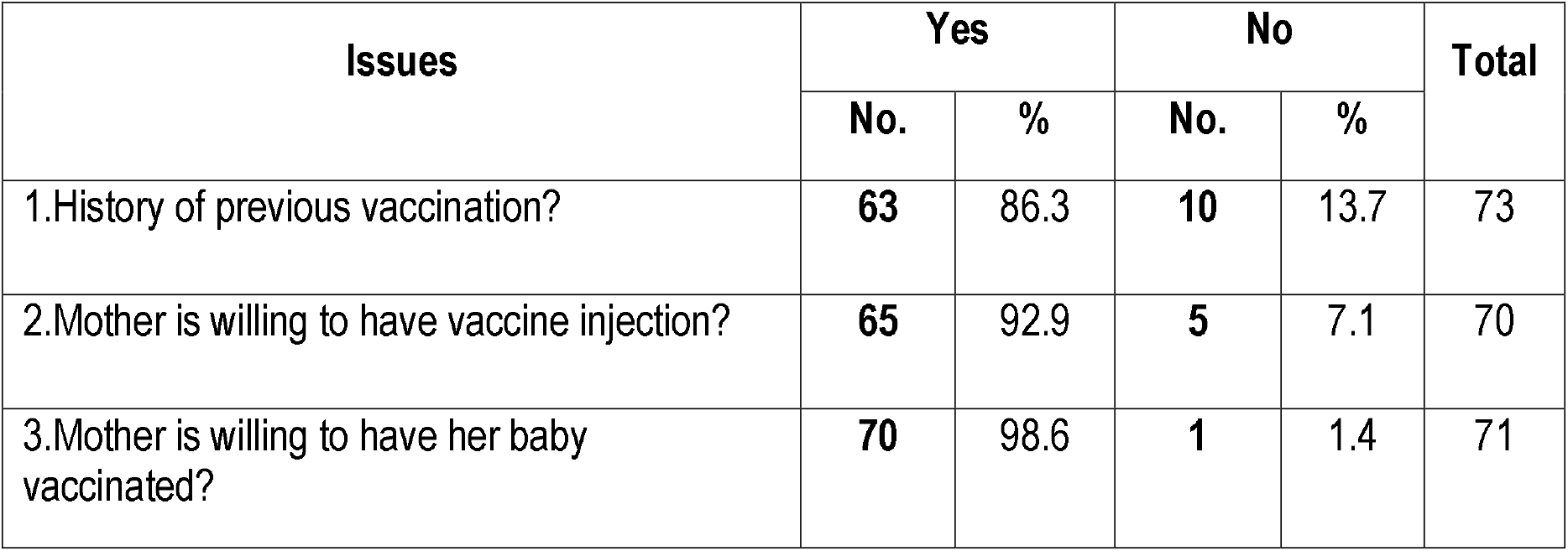
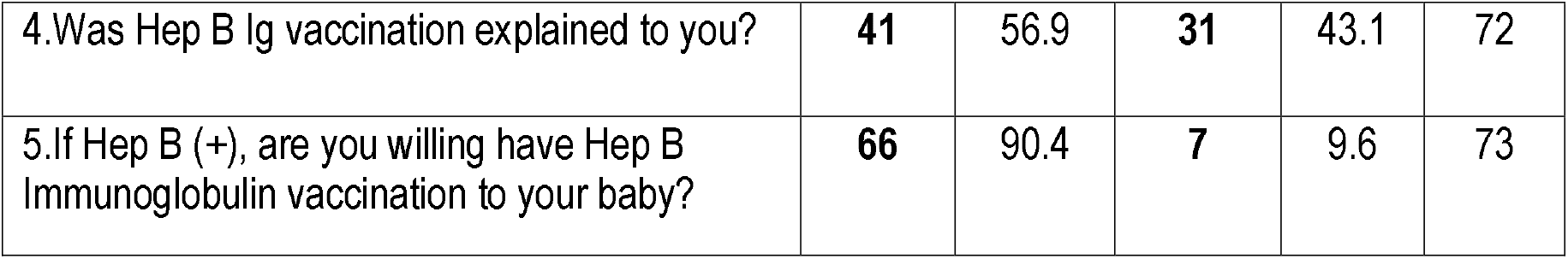
Knowledge of mothers on Hep B Ig vaccination(n=74).

| Issues | Yes |  | No |  | Total |
| --- | --- | --- | --- | --- | --- |
|  | No. | % | No. | % |  |
| 1.History of previous vaccination? | **63** | 86.3 | **10** | 13.7 | 73 |
| 2.Mother is willing to have vaccine injection? | **65** | 92.9 | **5** | 7.1 | 70 |
| 3.Mother is willing to have her baby vaccinated? | **70** | 98.6 | **1** | 1.4 | 71 |
| 4. Was Hep B Ig vaccination explained to you? | **41** | 56.9 | **31** | 43.1 | 72 |
| 5. If Hep B (+), are you willing have Hep B Immunoglobulin vaccination to your baby? | **66** | 90.4 | **7** | 9.6 | 73 |

Most of the respondents (48, 65.7%), preferred to have their immunizations at the barangay health centers, 50 (64.9%) indicated that vaccination should done by doctors, and followed, both by the nurses and midwives (17.1% each).

Regarding vaccination schedules, most of them preferred vaccination after their pregnancy period, 32 (44.4%). This data is presumed to be dependent on their knowledge after they were explained on hep B infection and hep B vaccine importance during their mothers’ classes. Other respondents would prefer vaccination before pregnancy (41.7%), and during breastfeeding (6.9%).

The least amount of vaccine at Php 1,550 was the choice by most of the respondents, 69 (97.2%). They will be paying on a 25% payment mode on every visit (19%) versus the full payment mode committed by only 9 mothers (11.4%) (Table 5). Moreover, the majority of them preferred those vaccines be funded from PHIC (44.3%) and be given for free.

**Table 5.** Preference of mothers on Hep B Ig vaccination(n=74).

| **Concerns** | **No.** | **%** | **Concerns** | **No.** | **%** |
| --- | --- | --- | --- | --- | --- |
| Facility preferred for vaccination | **73** |  | Preferred staff to do the injection | **74** |  |
| Brgy Health Center | 48 | 65.7 | Doctor | 50 | 64.9 |
| JRBGH | 25 | 34.2 | Nurse | 13 | 17.1 |
| Private Hospital | 0 | 0 | Midwife | 13 | 17.1 |
| Private Doctor | 0 | 0 |  |  |  |
| Price of Vaccine | **71** |  | Preferred timing of vaccination | **72** |  |
| Php 1,550 | 69 | 97.2 | Before pregnancy | 30 | 41.7 |
| Php 1,600 | 2 | 2.8 | After pregnancy | 32 | 44.4 |
| Php 1,650 | 0 | 0 | During breastfeeding | 5 | 6.9 |
| Php 1,700 | 0 | 0 | None | 5 | 6.9 |
| Php 1,750 | 0 | 0 |  |  |  |
| **Mode of Payment for the vaccine. Multiple answers. (n=79)** |  |  |  |  |  |
| Full payment | 9 | 11.4 | via PHIC | 35 | 44.3 |
| 50%-50% | 6 | 7.6 | via DOH | 9 | 11.4 |
| 25% per visit | 15 | 19.0 | Others | 3 | 3.8 |
| per Relatives | 2 | 2.5 |  |  |  |

Figure 2 shows the percentage of respondents who will recommend vaccination to other pregnant women. About 77.8% (56) recommended vaccination to other pregnant women. Only 2 (2.8%) will not recommend vaccination for any other reasons. Validation on these issues is beyond the scope of the study. There were 14 mothers who were not able to decide on whether to recommend vaccination or not (19.4%).

**Figure 2.**
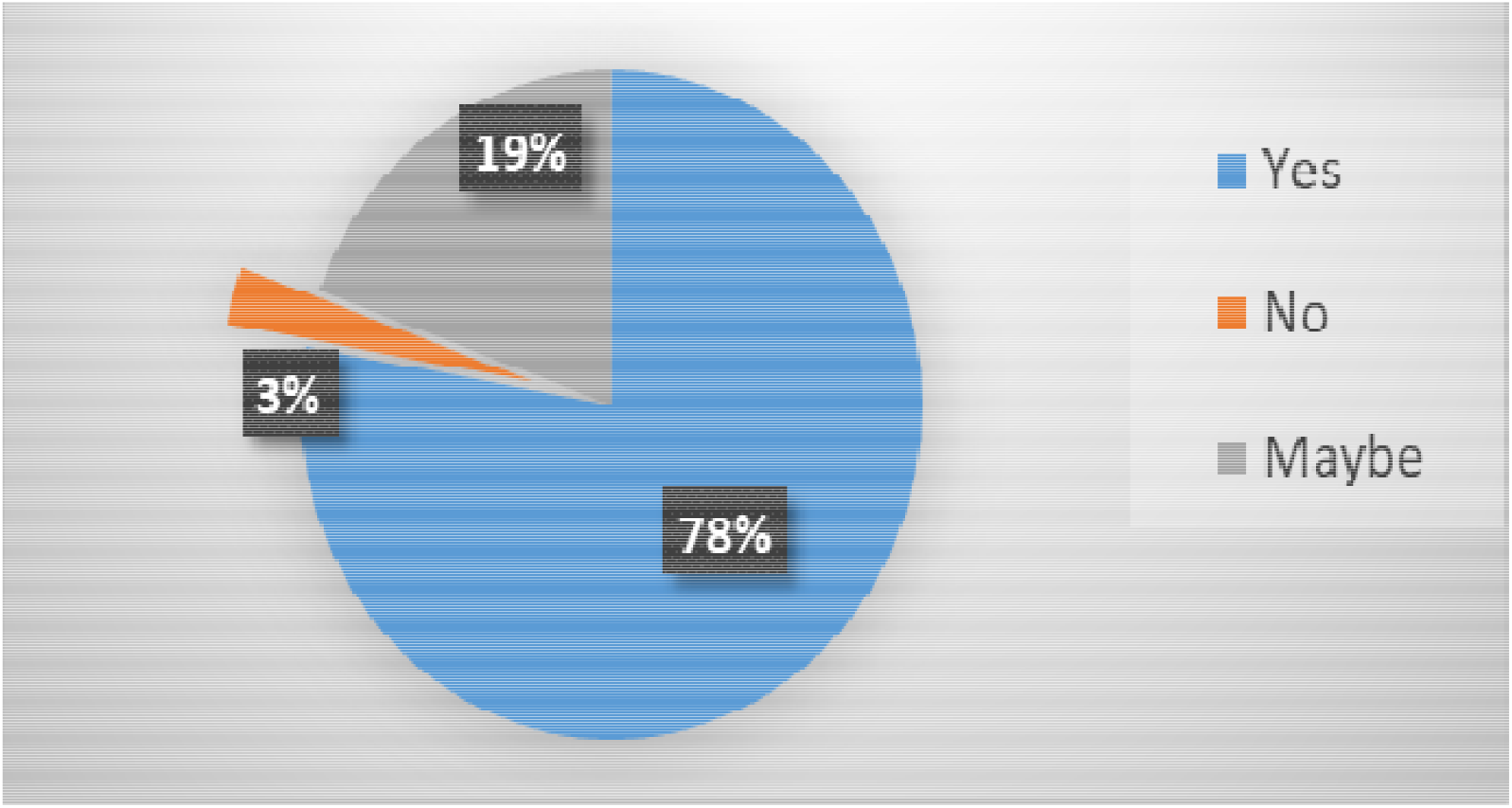
Percentage of mothers who will recommend vaccination to others.

The primary target market of the study were those babies born to hepatitis B reactive mothers (positive) during screening and who had their deliveries at the health facility. The majority of the respondents were PHIC members. Table 6 showed the total number of newborns born to pregnant women relative to the monthly total hospital admissions. Based on WHO (2017)^5^, hepatitis B infection estimated prevalence of 10% in the general population, there will be 2,404 newborns at high-risk for hepatitis B virus.

**Table 6.** JRBGH Population of patients seen, Obstetric Ward patients and Newborns for 2017.

| Months | Total Patients Seen | OB | % | Newborn | % | 10% Exposed |
| --- | --- | --- | --- | --- | --- | --- |
| Jan | 11621 | 4734 | 50.84 | **2310** | 19.88 | 231 |
| Feb | 9469 | 2999 | 38.86 | **1752** | 18.50 | 175.2 |
| Mar | 9006 | 3528 | 50.44 | **2011** | 22.33 | 201.1 |
| Apr | 11327 | 4141 | 45.58 | **2242** | 19.79 | 224.2 |
| May | 11199 | 3979 | 44.14 | **2184** | 19.50 | 218.4 |
| Jun | 10846 | 3751 | 42.91 | **2105** | 19.41 | 210.5 |
| Jul | 11231 | 3730 | 40.85 | **2101** | 18.71 | 210.1 |
| Aug | 12198 | 4098 | 41.42 | **2304** | 18.89 | 230.4 |
| Sep | 11420 | 4421 | 49.47 | **2484** | 21.75 | 248.4 |
| Oct | 10881 | 3892 | 44.72 | **2178** | 20.02 | 217.8 |
| Nov | 11160 | 3978 | 45.26 | **2371** | 21.25 | 237.1 |
| Dec | No data |  |  |  |  |  |
| **Total** | 120358 | 43251 |  | **24042** |  | 2404.2 |
| **Average** | 10,942 | 3,931 | 45% | 2,185 | 20% | 218 |

### B. SWOT Analysis

The SWOT analysis of the program is presented on Table 7. The internal strengths and weaknesses were shown as well as the external opportunities and possible threats which may affect in the sustainability of the program.

**Table 7.**
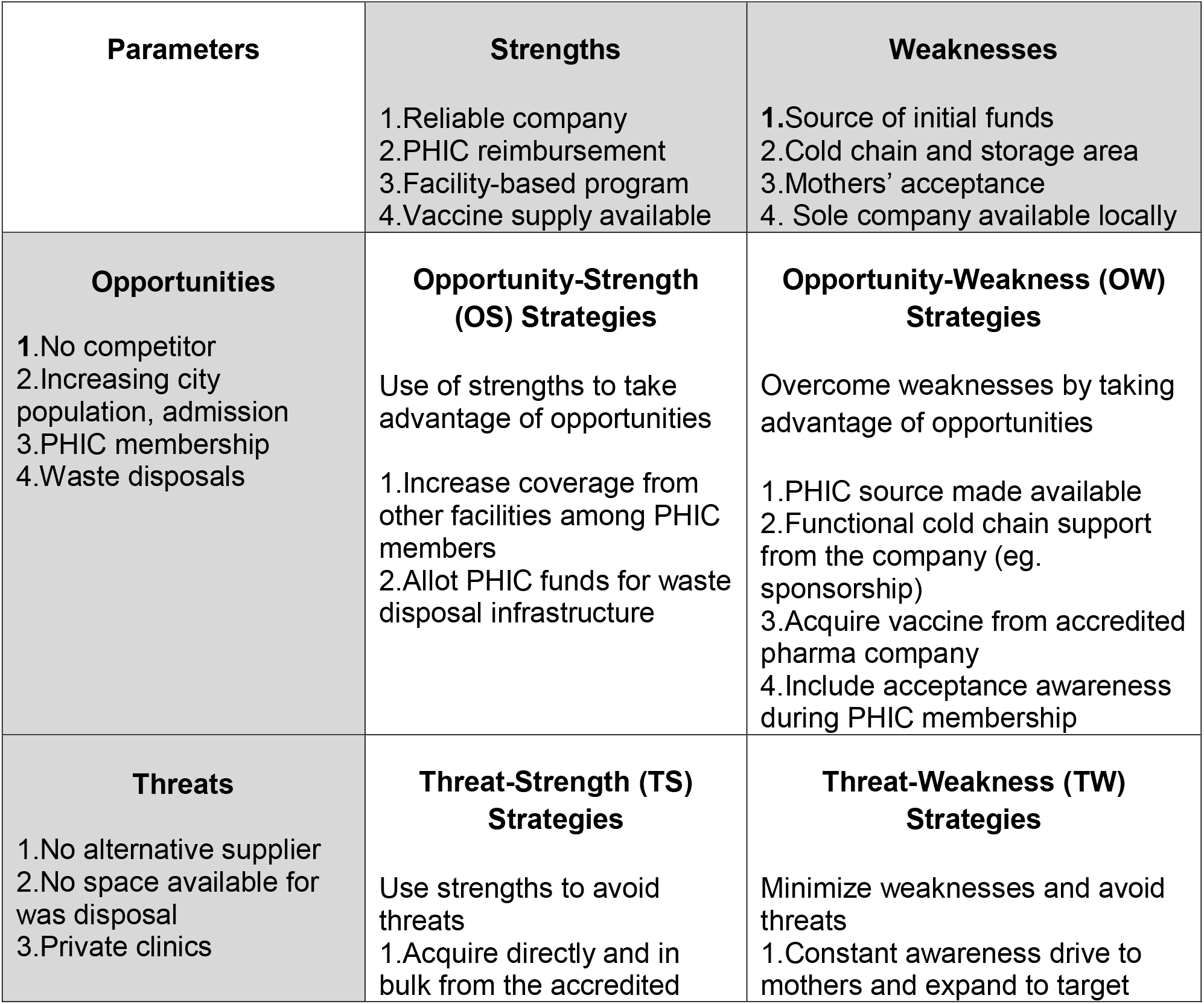
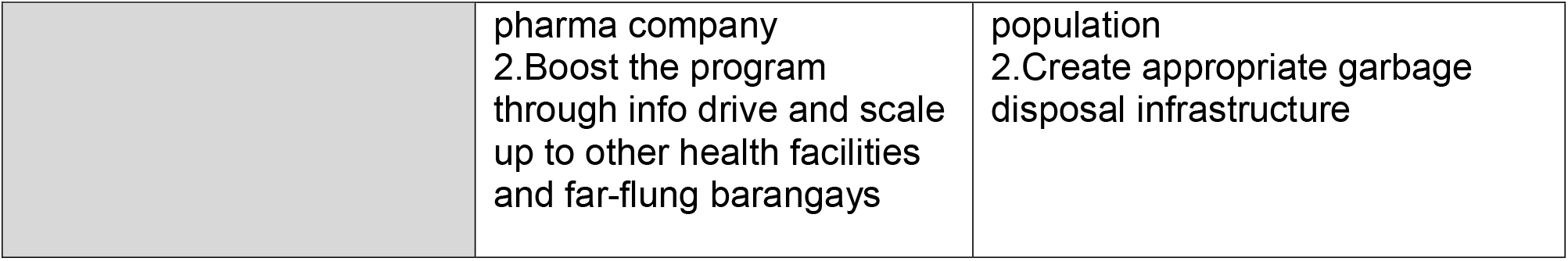
SWOT Grid on the proposed Hep B immunoglobulin vaccination program.

| **Parameters** | **Strengths** | **Weaknesses** |
| --- | --- | --- |
|  | 1.Reliable company<br>2.PHIC reimbursement<br>3.Facility-based program<br>4.Vaccine supply available | 1.Source of initial funds<br>2.Cold chain and storage area<br>3.Mothers' acceptance<br>4. Sole company available locally |
| **Opportunities** | **Opportunity-Strength (OS) Strategies** | **Opportunity-Weakness (OW) Strategies** |
| 1.No competitor<br>2.Increasing city population, admission<br>3.PHIC membership<br>4.Waste disposals | Use of strengths to take advantage of opportunities<br>1.Increase coverage from other facilities among PHIC members<br>2.Allot PHIC funds for waste disposal infrastructure | Overcome weaknesses by taking advantage of opportunities<br>1.PHIC source made available<br>2.Functional cold chain support from the company (eg. sponsorship)<br>3.Acquire vaccine from accredited pharma company<br>4.Include acceptance awareness during PHIC membership |
| **Threats** | **Threat-Strength (TS) Strategies** | **Threat-Weakness (TW) Strategies** |
| 1.No alternative supplier<br>2.No space available for was disposal<br>3.Private clinics | Use strengths to avoid threats<br>1.Acquire directly and in bulk from the accredited | Minimize weaknesses and avoid threats<br>1.Constant awareness drive to mothers and expand to target |
|  | pharma company<br>2.Boost the program through info drive and scale up to other health facilities and far-flung barangays | population<br>2.Create appropriate garbage disposal infrastructure |

### C. Financial Projections

The hospital average newborn deliveries were 2,185 babies with an average of 3,931 pregnant women gave birth per month (based on 2017 census, unpublished). If the WHO estimated Hepatitis B infection prevalence at 10%, then about 2,404 babies would be exposed to Hepatitis B virus from their mothers during deliveries or an average of 218 newborn per month. Moreover, the 2017 serology logbook from the hospital laboratory showed that the average HBsAG reactive mothers was 40 per month or 480 per year.

The financial 5-year projection was assumed based on the following: 3% increase in obstetrics admission, 480 hep B-reactive mothers per year (2017), national health insurance reimbursement (PHIC) at Php 4,000, and the amount of Php 1,550.00 as the preferred amount for Hep B Ig vaccine (single dose only) (Table 8). Prevailing dollar exchange average at Php 51.7675= 1 dollar at the time of study period. The projected revenue would total to Php 1,494,500.00 ($28,869.5) on the 5^th^ year and would spare the mothers from OOP expenses with a total amount of Php 945,500 ($18,264.40).

**Table 8.** Five-year forecast for the revenue of Hep B Ig vaccination among newborns born to Hepatitis-B reactive mothers at JRBGH.

|  | Parameters | 2016 | 2017 | 2018 | 2019 | 2020 | 2021 |
| --- | --- | --- | --- | --- | --- | --- | --- |
| A | OB cases (3%) | 6,715 | 6,915 | 7,122 | 7,336 | 7,556 | 7,782 |
| B | Projected HepB(+)/yr (eg. 480 x 7%) | - | 480 | 498 | 533 | 570 | 610 |
| C | Difference (yearly) (eg. 2018-2017) | - | - | 18 | 35 | 37 | 40 |
| D | Amt of vaccine (B x 1550) | - | 744,000 | 771,900 | 826,150 | 883,500 | 945,500 |
| E | PHIC per Hep B Ig (B x 4,000) | - | 1,920,000 | 1,992,000 | 2,132,000 | 2,280,000 | 2,440,000 |
| F | Revenue (E-D) | - | **1,176,000** | **1,220,100** | **1,305,850** | **1,396,500** | **1,494,500** |
| G | Increment (yearly) (eg.2018-2017) | - |  | 44,100 | 85,750 | 90,650 | 98,000 |

**Table 9.** Projected monthly forecast for the revenue of Hep B Ig vaccination among newborns born to Hepatitis B reactive mothers in Cagayan De Oro City (based on 2016 census).

| Months | Newborn Deliveries | Hep B (+) | Amt of Hep B vaccine* | PHIC reimbursement* | Revenue* | Difference* |
| --- | --- | --- | --- | --- | --- | --- |
| Jan | **1354** | 95 | 146,909 | 379,120 | **232,211** | - |
| Feb | **1184** | 83 | 128,464 | 331,520 | **203,056** | -29,155 |
| March | **1420** | 99 | 154,070 | 397,600 | **243,530** | 40,474 |
| April | **1553** | 109 | 168,501 | 434,840 | **266,340** | 22,810 |
| May | **1710** | 120 | 185,535 | 478,800 | **293,265** | 26,925 |
| June | **1537** | 108 | 166,765 | 430,360 | **263,596** | -29,670 |
| July | **1442** | 101 | 156,457 | 403,760 | **247,303** | -16,293 |
| Aug | **1621** | 113 | 175,879 | 453,880 | **278,002** | 30,699 |
| Sept | **1874** | 131 | 203,329 | 524,720 | **321,391** | 43,390 |
| Oct | **1619** | 113 | 175,662 | 453,320 | **277,659** | -43,733 |
| Nov | **2010** | 141 | 218,085 | 562,800 | **344,715** | 67,057 |
| Dec | **2391** | 167 | 259,424 | 669,480 | **410,057** | 65,341 |
| **Total** | **19,715** | **1,380** | **2,139,078** | **5,520,200** | **3,381,123**<br>(Ave.=281.760.25) |  |

Note: \* = in pesos

Moreover, Table 8 showed the monthly revenue of Hep B Ig vaccination (2016 census) on a city-wide scale based on the following assumptions: 40 hep B-reactive mothers per month; 480 hep B-reactive mothers or 7% of the 6,915 obstetric cases (2017); Php 4,000 as the reimbursable amount by PHIC for hep B Ig vaccination; and Php 1,550 as the prevailing and preferred cost of hep B Ig vaccine. Based on the above assumptions, there were a total of 1.380 newborns that would be at risk to hepatitis B-reactive mothers. The difference of the total amount of PHIC reimbursement from the amount of vaccine purchasing would give the facility a projected revenue of Php 3,381,123.00 ($65,313.62) or a monthly average of Php (281,760.25).

## DISCUSSION

Clinically, hep B immunoglobulin is highly recommended to protect the newborn population against mother to child via vertical transmission of hepatitis B virus. It is also highly recommended to give the vaccine within 12 hours of life. This target population, as they become an adult, would carry these protective measures against future cohort of hepatitis B infected individuals. The local government and the healthcare institutions shall be in position to make these vaccines accessible since it is not part of the government’s EPI program. Locally, there is no published and available study assessing the health cost and burden of hepatitis B Ig in a government-owned hospital. For decision-making, empirical evidence are very relevant and logical. Financial managers should deal in-depth economic studies for future and current use, an da s baseline study for scaling u the program to a national level.

Other studies pointed out cost-effective measures to incorporate hepatitis B immunoglobulin with hepatitis B primary vaccination and screening pregnant women^6^, and or hepatitis B vaccination combined with immunoglobulin^7^. This current study contributes to evidence-based understanding of the financial burden of delivering the Hep B Ig. Meanwhile, the healthcare facility will proactively procure the vaccine with the available resources.

This documented the acceptance of hep B Ig on the demand side of a vaccine program. If prospectively conducted for the 2,404 newborns, this target population may contribute to the rising number of complicated or non-complicated chronic hepatitis B infection. We must help these mothers to make informed decisions for the hep B Ig immunization (and for the other vaccines) through daily prenatal/ mothers’ class, family planning counselling, the postpartum classes for better acceptance of the vaccine as a program, compliance and wider coverage. Implementation of universal hepatitis B vaccination in the country should still be pushed forward for no newborn should be left behind. Optimal national universal hepatitis vaccination requires extensive efforts to overcome current social and economic challenges. Hepatitis B immunoglobulin vaccine should be included in these programs.

The burden of its cost is way behind the cost of chronic hepatitis treatment. Furthermore, mortality due to viral hepatitis is estimated at 29% due to cirrhosis secondary to viral hepatitis, 3% due to acute hepatitis, and 67% due to liver cancer^8^. All are preventable and successfully reduced with the maximum implementation of hepatitis vaccination programs.

On the supply side of the vaccination program, hep B Ig vaccine should be made available 24 hours and 7 days a week. Currently these are available in private clinics, hospitals and pharmacies around the city. This study presented a financial 5-year forecast with a good revenue amounting to Php 1,494,500.00 ($28,869.5) on its 5^th^ year and would spare the mothers from unnecessary out-of-pocket expenditures amounting to Php 945,500 ($18,264.40) or almost a million pesos.

The study presented the cost of Hep B Ig for the beneficiaries and for the facilities handling the Hepatitis B exposed newborns. As a preventable disease, maternal awareness and knowledge of the vaccine at the first facility visits will greatly expand the cohort of newborn with the Hep B Ig coverage (output), expected outcome will reduce hepatitis and liver cancer, and sustainably impact towards a reduced health burden of Hepatitis B infection (impact).

The prevailing cost is dependent on the prevailing world market and may change overtime or within the timeframe of the cost forecast. Moreover, projected city-wide population may fluctuate as well as the availability of the service per se especially during pandemic.

Initial engagement with the facility were more on discussions based on the situational analysis and the clinical needs of the newborn. Hep B Ig was purchased in bulk at the health facility’s pharmacy, accessible by the mothers anytime for in-patients, and newborns were given Hep B Ig within the critical period of 24 hours without out-of-pocket expenditures.

This study had several strengths and limitations. Four limitations were identified: 1. Published census data for the city were outdated almost 4 years back, and not current as to the time of study period, 2. Health costing mainly utilized hepatitis B immunoglobulin vaccine and did not include the supplies and other materials during vaccination (eg. Cotton balls), 3. Small number of respondents was used in the study and recommended for a much larger group, 4. Newborn hepatitis b exposure were determined sole using the HBsAg titer among the screened pregnant women.

However, this study presented a baseline data for hepatitis B Ig cost analysis, had random respondents representing each top ten barangays, and hospitals and laboratory data were collated and updated.

The protective measures from vaccination still high and the demand is increasing. It is still considered as the primary preventive measures against viral hepatitis in a larger scale. Furthermore, considered also as a very cost-effective preventive health measure to date. The local government’s advocacy for Health for All is well supported and hopefully

Hepatitis B immunoglobulin vaccine will be part of it nationally. As the healthcare industry continues to have costs spiraling upwards, the majority of the city residents cannot afford the vaccination outside of the Expanded Program of Immunization program.

These newborns will be best supported by the wider and extensive coverage under the Universal Health Care of the national government to reduce the economic risk of out-of-pocket expenses among pregnant women and more protection among the younger generations.

## Data Availability

All data produced in the present work are contained in the manuscript

## OTHER RELEVANT INFORMATION

### Reporting Guidelines

This study utilizes the Enhancing the QUAlity and Transparency Of Health Research^9^ upon submission.

